# Performance evaluation of TaqMan^™^ Arbovirus Triplex Kit (ZIKV/DENV/CHIKV) for detection and differentiation of Dengue and Chikungunya viral RNA in serum samples of symptomatic patients

**DOI:** 10.1101/2024.06.12.24308802

**Authors:** Kakhangchung Panmei, Syed Abdul Hakeem, Obiageli Okafor, Shoba Mammen, Asha Mary Abraham

## Abstract

**Introduction:** Global outbreaks of mosquito-transmitted arbovirus infections, such as dengue (DENV) and chikungunya (CHIKV), are increasing. Differentiating these infections is challenging due to non-specific symptoms and serology limitations. PCR-based approaches offer higher sensitivity and specificity. This study evaluated the performance of TaqMan™ Arbovirus Triplex Kit (ZIKV/DENV/CHIKV) (TaqMan™ Kit) to detect DENV and CHIKV in clinical samples from patients in south India.

**Methods:** In total, 280 serum samples with 90 DENV-positive, 90 CHIKV-positive, and 100 negative samples were tested with TaqMan™ Kit and CDC Trioplex Real-Time RT-PCR assay. No Zika virus was detected. The sensitivity and specificity of viral RNA detection were determined, and discordant results were resolved using comparator PCRs, dengue NS1 antigen detection, virus-specific antibody results, or previously de-identified in-house PCR results.

**Results:** The TaqMan™ Kit showed 100% agreement with the comparator for DENV detection in 92 positive samples. Among 188 samples negative for DENV by the comparator, 30 showed positive results with the TaqMan™ kit, and 23 of those were confirmed as true positives. The resulting sensitivity and specificity for DENV detection were 100% and 95.1%, respectively. For CHIKV, 77 positive and 195 negative results were concordant. Eight samples showed discordant results, but upon resolution testing, sensitivity and specificity for CHIKV were 93.9% and 100.0%, respectively.

**Conclusion:** The TaqMan™ Arbovirus Triplex Kit demonstrated high sensitivity and specificity (>93%) for detecting circulating DENV and CHIKV strains. Multiplex PCR testing can improve case detection, surveillance, and public health responses while optimizing laboratory resources for outbreak control.

## Introduction

Arboviral diseases, transmitted to humans by bites of infected arthropods, mainly mosquitoes, represent a substantial global public health problem (1). Dengue (DENV) and chikungunya (CHIKV) viruses are arboviruses that are transmitted by mosquitoes of the *Aedes spp*., with *A. aegypti* and *A. albopictus* considered to be the main vectors (2). *A. aegypti* and *A. albopictus* currently have wide distribution, covering all continents (except Antarctic), though mainly found in tropic and subtropic regions, where the burden of arboviral disease is the highest (3–5). Dengue has the highest prevalence among the arbovirus infections. According to the World Health Organization (WHO), over 390 million dengue virus infections occur each year, with up to half of the world’s population at risk of being infected (6).

India has experienced a significant increase in arbovirus infections in recent years. Reported cases of dengue doubled from approximately 100,000 cases in 2018 to over 230,000 cases in 2022 and dengue-related deaths increased by 76% (7). During this period, suspected cases of chikungunya in India also experienced a remarkable increase of over 150% in less than 5 years (8). Beyond the impact on individual health, arboviral diseases burden healthcare systems and national economies. For instance, estimated productivity losses during the chikungunya epidemic in 2016 ranged between INR 214.4 and 391 million per year (equivalent to USD 2.57–4.69 million) (9). For the same year (2016), a study estimated the total indirect costs associated with dengue, including the economic output foregone due to illness and premature death, to be USD 1,607,159,023. The study also revealed that the cost of a hospitalized dengue case was approximately 20 times higher than that of a non-medical case, and a fatal dengue case was about 100 times higher in cost than a hospitalized case (10). A similar study in 2018, observed varying median direct costs, among different severity levels of pediatric and adult dengue cases. For pediatric dengue, the median direct costs were 179.80 USD for cases without warning signs, 145.06 USD for cases with warning signs, and 933.51 USD for severe cases., Conversely for adult dengue, the median direct costs were 312.75 USD, 287.22 USD, and 720.39 USD for cases without warning signs, with warning signs, and severe cases, respectively. Healthcare expenses, including treatment and indirect costs, significantly impact many Indians, pushing an estimated 55 million into poverty every year (11).

The true extent of the disease burden may be significantly underestimated due to inadequate and inaccurate reporting. Co-circulation of multiple Aedes-transmitted diseases with similar epidemiology and often clinically indistinguishable febrile syndromes make diagnosis challenging in the absence of reliable tests. Both antibody detection tests such as enzyme linked immuno-assays (ELISA), antigen detection ELISA’s and molecular tests (PCR) have been widely used for arbovirus detection. However, the phase of infection at time of sample collection or exposure to other antigenically related viruses can affect the result validity (12,13). Both DENV and CHIKV can be detected in the blood as early as 2 days before the onset of symptoms. The viral load typically peaks around the onset of clinical symptoms and gradually declines towards the end of the initial week of clinical illness just as viral antibodies start to develop (14,15). As a result, serological tests such as immunoglobulin M (IgM) ELISA and even neutralization assays are unable to confirm acute phase infections. In addition, immuno-assays may not reliably identify the infecting virus due to previous exposure to related flaviviruses and extensive cross-reactivity among flaviviruses (16). NS1 antigen ELISAs and PCR can be used during this phase. However, commercial PCRs are not readily available and may require additional facilities for setup.

Current WHO guidance on laboratory testing for DENV and CHIKV recommends the use of molecular assays as the preferred method over serological tests for acute-phase infections (17,18). The main advantages of PCR testing are high sensitivity and specificity and the ability to detect acute-phase infections. Employing the most suitable test at the appropriate stage of infection allows for early detection and is vital for effective case detection and surveillance of arboviruses for monitoring of disease trends, early identification of outbreaks, and implementation of targeted public health interventions (19).

As arboviruses are RNA viruses that have a high mutation rate, evaluations of available tests for the detection of these viruses on recently circulating strains are important to ensure reliable results. Access to accurate testing modalities that can ensure early, highly sensitive and specific detection and differentiation of arboviruses is a crucial element in implementing effective surveillance programs. This study aims to evaluate the performance of the Research Use Only (RUO) TaqMan™ Arbovirus Triplex Kit (ZIKV/DENV/CHIKV) (TaqMan™ Arbovirus kit) to detect circulating strains of DENV and CHIKV in clinical samples of patients from a tertiary care hospital in south India.

### Methodology

This study was designed as a retrospective cross-sectional analytical study from archived serum samples previously collected from symptomatic patients (of all ages) by a healthcare provider. The study was conducted in Christian Medical College, Vellore (CMC) and was approved by the Institutional Research and Ethics Committee (IRB Min.No. 14750 [DIAGNO], dated 27.7.2022).

Serum samples were collected (convenient sampling) from the department of Clinical Virology, CMC Vellore. 2,545 samples were screened using an in-house PCR for Zika virus (ZIKV), CHIKV & DENV of which 280 samples were identified for testing (90 positives for DENV and CHIKV and 100 negatives for all three targets i.e., DENV, CHIKV and ZIKV). The PCR positive samples recruited for DENV & CHIKV were with or without serology results. There were no ZIKV positive samples. All samples after inclusion were de-identified and given a unique study ID. Templates for nucleic acid extraction and PCR were made by random sampling. For every batch of samples extracted, testing with TaqMan™ Arbovirus kit and Trioplex Real-time RT-PCR assay (CDC Trioplex) were carried out on the same day

### Nucleic acid extraction

For each sample, 200μl of serum was used for extraction (as per the user the manufacturers guideline) using the Qiagen QIAamp 96 Virus QIAcube HT kit automated platform (20).

### Testing method-1 (TaqMan™ Arbovirus kit)

The TaqMan™ Arbovirus kit (RUO) comes in a lyophilized form of primers and probes for viral (ZIKV, CHIKV, DENV) and internal control - Peptidylprolyl Isomerase A (PPIA) targets in a 0.1 mL sample PCR well. Using the Applied Biosystems™ QuantStudio™ 6 Flex (QS6) Real-Time PCR System, each sample was tested as a single multiplex rtRT-PCR reaction. Each test utilized a 25µL elution volume and followed a run method consisting of 50 °C for 20 minutes, 95 °C for 2 minutes, 45 cycles at 95 °C for 15 seconds, and 60 °C for 1 minute (acquisition)(21).

### Comparator-1 (CDC Trioplex)

CDC Trioplex was used as a comparator. The samples were tested in a two-tube-25µL PCR reaction, one tube multiplex rtRT-PCR for ZIKV, CHIKV, DENV and another for internal control (RNAse P). It was performed on QS6 Real-Time PCR system, using 15µL PCR buffer plus 10µL sample elution volume in 0.1 mL sample PCR wells at 50 °C for 30 mins, 95 °C for 2 minutes, 45 cycles at 95 °C for 15 secs and 60 °C 1 min (acquisition) (as per the kit protocol)(22).

Results were analyzed according to the standard laboratory and published assay validation protocols and manufacturer’s instructions for CDC Trioplex and TaqMan™ Arbovirus Kit with the accompanying instrument software (21–23). A Ct value of ≤38 was considered positive while >38 was considered negative for both assays. All the DENV-positive samples, regardless of the assay used, were further analyzed for DENV-specific serotype using CDC DENV-1-4 rRT-PCR assay (23). The discordant samples were resolved based on three categories (Fig 1).

**Fig 1:**
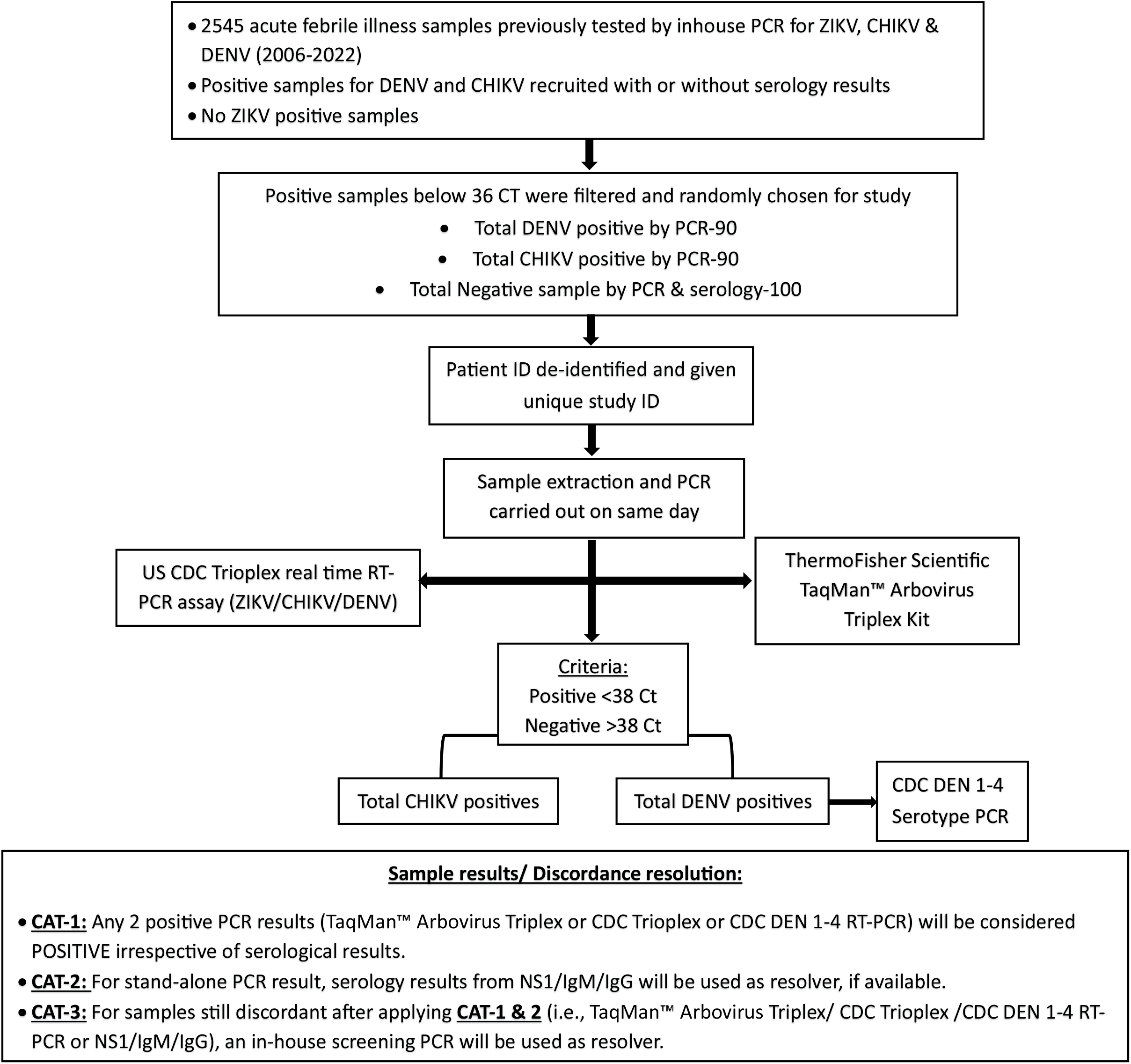
Flowchart of the methodology

### Resolver CAT-1 (CDC DENV-1-4 rRT-PCR)

#### DENV discordant samples

Samples that tested positive for DENV using the TaqMan™ Arbovirus kit but negative with the CDC Trioplex assay, and vice versa, underwent further analysis using DENV serotype-specific PCR i.e., CDC DENV-1-4 rRT-PCR assay to resolve discrepancies (Fig 2).

**Fig 2:**
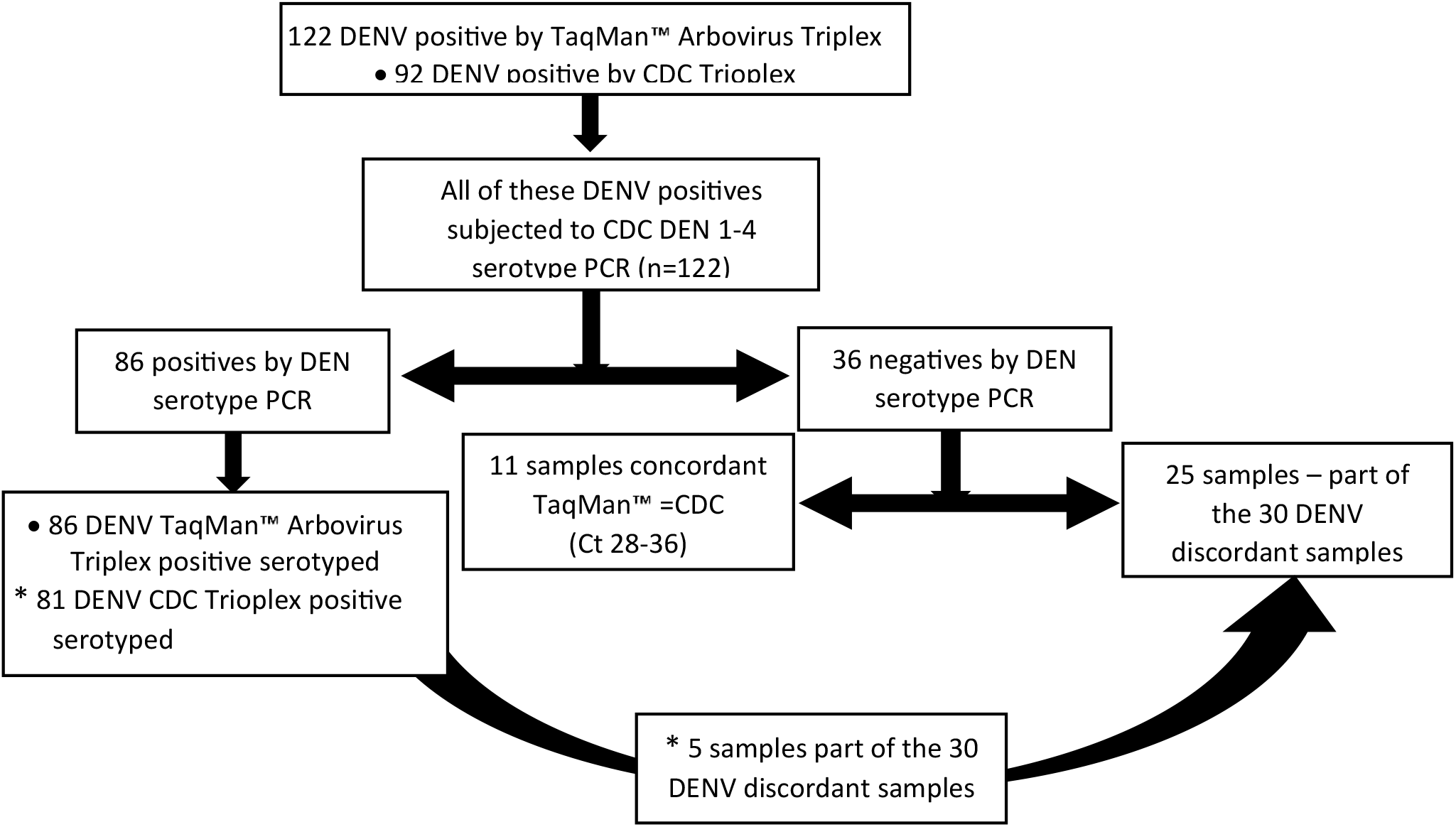
Flowchart of DENV positives subjected to CDC DENV 1-4 rRT-PCR

### Resolver CAT-2 (DENV NS1, IgM & IgG & CHIKV IgM capture ELISA)

#### CHIKV discordant samples

Discordant samples that were positive for CHIKV in only one of the assays were subjected to serology with CHIKV IgM ELISA for resolution.

#### DENV discordant samples

DENV samples that remained discordant after applying resolver-1 were further analyzed for evidence of viral infection using serology i.e., DENV NS1, IgM & IgG capture ELISA.

### Resolver CAT-3 (Screening RT-PCR)

To achieve a final resolution of discordant samples that remained even after applying resolvers 1 & 2, the in-house screening RT-PCR method initially used to select samples for this study was employed.

## Results

### CHIKV pre-resolution results

Of the 280 samples tested using both multiplex assays for ZIKV, CHIKV, and DENV, the TaqMan™ Arbovirus kit detected a total of 78 CHIKV positives and 202 CHIKV negatives while CDC Trioplex identified a total of 84 CHIKV positives and 196 CHIKV negatives. There were 77 and 195 concordant samples for CHIKV positives and negatives respectively **(Supplement Table 1a, 1b)**. Eight samples were discordant, of which 7 samples negative by TaqMan™ Arbovirus kit were positive by CDC Trioplex while 1 sample positive by TaqMan™ Arbovirus kit was negative by CDC Trioplex. Positive percent agreement (PPA) and negative percent agreement (NPA) were 91.67% and 99.49% respectively. The accuracy of the assay was 97.14 % with a Kappa coefficient (CI-95%) agreement of 0.93 **(Table 1)**.

**Table 1.** Pre-resolution agreements of TaqMan™ Arbovirus kit with CDC Trioplex for CHIKV.

| TaqMan™ Arbovirus Triplex | CDC Trioplex |  |  |  |
| --- | --- | --- | --- | --- |
|  |  | + | - | Total |
|  | + | 77 | 1 | 78 |
|  | - | 7 | 195 | 202 |
|  | Total | 84 | 196 | 280 |
| PPA (CI 95%) | 91.67 (83.58 - 96.58) |  |  |  |
| NPA (CI 95%) | 99.49 (97.19 - 99.99) |  |  |  |
| Accuracy (%) | 97.14 |  |  |  |
| Kappa Coefficient (CI 95%) | 0.93 (0.88 - 0.98) |  |  |  |

Note. PPA, positive percent agreement; NPA, negative percent agreement.

### CHIKV post-resolution results

Cat-2 and Cat-3 resolved all eight discordant CHIKV samples. Among these, five of seven samples initially negative by TaqMan™ Arbovirus kit but positive by CDC Trioplex were resolved as positive for CHIKV and 2 samples as CHIKV negative. The remaining discordant sample (ZVT-93) which was negative on the CDC trioplex assay but with a Ct value of 33.21 on the TaqMan™ Arbovirus kit was resolved as CHIKV positive consistent with TaqMan™ Arbovirus results **(Supplement Table 2)**. Upon resolution, the sensitivity, and specificity (CI-95%) marginally increase to 93.98% and 100% with a PPV and NPV of 100% and 97.52% respectively. The overall accuracy increased slightly to 98.21% and Kappa coefficient (CI-95%) agreement to 0.96 **(Table 2)**.

**Table 2.** Performance of TaqMan™ Arbovirus kit for CHIKV.

| **TaqMan™ Arbovirus Triplex** | **CDC Trioplex** |  |  |  |
| --- | --- | --- | --- | --- |
|  |  | + | - | Total |
|  | + | 78 | 0 | 78 |
|  | - | 5 | 197 | 202 |
|  | Total | 83 | 197 | 280 |
| Sensitivity (CI 95%) | 93.98 (86.50 - 98.02) |  |  |  |
| Specificity (CI 95%) | 100.00 (98.14- 100.00) |  |  |  |
| Accuracy (%) | 98.21 |  |  |  |
| PPV (%) | 100.00 |  |  |  |
| NPV (%) | 97.52 |  |  |  |
| Kappa Coefficient (CI 95%) | 0.96 (0.92 - 0.99) |  |  |  |

Note. PPV, positive predictive value; NPV, negative predictive value.

### DENV pre-resolution results

TaqMan™ Arbovirus kit identified 122 DENV positives and 158 DENV negatives, while CDC Trioplex assay detected 92 DENV positives and 188 DENV negatives. There were 92 and 158 DENV concordant positives and negatives respectively **(Supplement Table 3a, 3b)**. The PPA and NPA were 100% and 84.04% respectively. The overall accuracy of the assay was 89.29 % with a Kappa coefficient (CI-95%) agreement of 0.78 **(Table 3)**. For resolution, the 122 DENV positives were subjected to CDC DENV-1-4 rRT-PCR of which 86 and 81 samples positive by TaqMan™ Arbovirus kit and by CDC Trioplex were serotyped **(Supplement Table 4)**. 30 samples showed discordant results for DENV, testing positive with the TaqMan™ Arbovirus kit and negative with the CDC Trioplex **(Table 4)**.

**Table 3.** Pre-resolution agreements of TaqMan™ Arbovirus kit with CDC Trioplex for DENV.

| **TaqMan™ Arbovirus Triplex** | **CDC Trioplex** |  |  |  |
| --- | --- | --- | --- | --- |
|  |  | + | - | Total |
|  | + | 92 | 30 | 122 |
|  | - | 0 | 158 | 158 |
|  | Total | 92 | 188 | 280 |
| PPA (CI 95%) | 100.00 (96.07 – 100.00) |  |  |  |
| NPA (CI 95%) | 84.04 (78.01 - 88.97) |  |  |  |
| Accuracy (%) | 89.29 |  |  |  |
| Kappa Coefficient (CI 95%) | 0.78 (0.70 - 0.85) |  |  |  |

Note. PPA, positive percent agreement; NPA, negative percent agreement.

**Table 4.** DENV discordant samples.

| S.No | Study ID | TaqMan™ (DENV Ct value) | CDC Trioplex DENV Ct value | DENV Serology | CDC DENV 1-4 Serotype | TaqMan™ CHIKV Ct value | CDC Trioplex CHIKV Ct value | DENV post resolution results (resolved by) |
| --- | --- | --- | --- | --- | --- | --- | --- | --- |
| 1 | ZVT-103 | Positive (29.12) | 0 | Positive | Negative | 0 | 0 | DENV positive (Cat-2) |
| 2 | ZVT-104 | Positive (29.10) | 0 | Positive | Negative | 0 | 0 | DENV positive (Cat-2) |
| 3 | ZVT-115 | Positive (35.79) | 0 | Positive | Negative | 0 | 0 | DENV positive (Cat-2) |
| 4 | ZVT-116 | Positive (35.62) | 0 | Positive | Negative | 0 | 0 | DENV positive (Cat-2) |
| 5 | ZVT-120 | Positive (28.58) | 0 | Positive | Negative | 0 | 0 | DENV positive (Cat-2) |
| 6 | ZVT-122 | Positive (34.03) | 0 | Positive | Negative | 0 | 0 | DENV positive (Cat-2) |
| 7 | ZVT-137 | Positive (36.81) | 39.01<sup>#</sup> | Negative | Negative | 0 | 0 | Negative (Cat-2) |
| 8 | ZVT-146 | Positive (30.70) | 0 | Positive | DEN-3 | 0 | 0 | DENV positive (Cat-1 & 2) |
| 9 | ZVT-227 | Positive (28.63) | 0 | Positive | Negative | 0 | 0 | DENV positive (Cat-2) |
| 10 | ZVT-157 | Positive (31.49) | 0 | Positive | Negative | 0 | 0 | DENV positive (Cat-2) |
| 11 | ZVT-161 | Positive (34.04) | 0 | Positive | Negative | 0 | 0 | DENV positive (Cat-2) |
| 12 | ZVT-165 | Positive (33.56) | 0 | Positive | DEN-1 | 0 | 0 | DENV positive (Cat-1 & 2) |
| 13 | ZVT-09* | Positive (35.98) | 0 | Negative | Negative | 12.38 | 16.39 | CHIKV positive (Cat-2) |
| 14 | ZVT-168 | Positive (35.21) | 0 | Positive | Negative | 0 | 0 | DENV positive (Cat-2) |
| 15 | ZVT-169 | Positive (36.39) | 0 | Positive | Negative | 0 | 0 | DENV positive (Cat-2) |
| 16 | ZVT-170 | Positive (32.26) | 0 | Positive | Negative | 0 | 0 | DENV positive (Cat-2) |
| 17 | ZVT-171 | Positive (30.62) | 0 | Positive | Negative | 0 | 0 | DENV positive (Cat-2) |
| 18 | ZVT-172 | Positive (31.46) | 0 | Positive | Negative | 0 | 0 | DENV positive (Cat-2) |
| 19 | ZVT-173 | Positive (36.30) | 0 | Positive | Negative | 0 | 0 | DENV positive (Cat-2) |
| 20 | ZVT-238 | Positive (29.56) | 0 | Positive | Negative | 0 | 0 | DENV positive (Cat-2) |
| 21 | ZVT-240 | Positive (29.82) | 0 | Positive | DEN-2 | 0 | 0 | DENV positive (Cat-1 & 2) |
| 22 | ZVT-25* | Positive (36.14) | 0 | Negative | Negative | 17.82 | 22.33 | CHIKV positive (Cat-2) |
| 23 | ZVT-26* | Positive (36.89) | 0 | Negative | Negative | 19.43 | 23.71 | CHIKV positive (Cat-2) |
| 24 | ZVT-52* | Positive (35.93) | 0 | Negative | Negative | 23.09 | 26.98 | CHIKV positive (Cat-2) |
| 25 | ZVT-53* | Positive (36.97) | 0 | Negative | Negative | 22.29 | 26.08 | CHIKV positive (Cat-2) |
| 26 | ZVT-54* | Positive (35.92) | 0 | Negative | Negative | 21.27 | 25.48 | CHIKV positive (Cat-2) |
| 27 | ZVT-190 | Positive (35.91) | 0 | Positive | Negative | 0 | 0 | DENV positive (Cat-2) |
| 28 | ZVT-252 | Positive (30.69) | 0 | Positive | DEN-2 | 0 | 0 | DENV positive (Cat-1 & 2) |
| 29 | ZVT-253 | Positive (30.81) | 0 | Positive | DEN-2 | 0 | 0 | DENV positive (Cat-1 & 2) |
| 30 | ZVT-28* | Positive (35.17) | 0 | Negative | Negative | 21.29 | 24.83 | CHIKV positive (Cat-2) |

### DENV post-resolution results

All 30 discordant DENV samples positive only by TaqMan™ Arbovirus kit PCR were resolved by **Cat-1 and Cat-2 (Table 4)**. 5 samples were positive for DENV by serology and serotype PCR, while 17 samples were positive for DENV by serology only. Eight samples with low CHIKV Ct values showed co-positivity with DENV at high Ct values in the TaqMan™ Arbovirus kit **(Table 5)**. All 8 samples were DENV-negative by Cat-2 as DENV NS1 and IgM/IgG antibodies were absent but for one sample, both TaqMan™ Arbovirus kit and CDC Trioplex confirmed co-positivity. Post resolution analysis for sensitivity, and specificity (CI-95%) stood at 100% and 95.18 % respectively. The PPV was 93.44% and NPV was 97.52% respectively. The overall accuracy increased to 97.14% with a Kappa coefficient (CI-95%) agreement of 0.94 **(Table 6)**.

**Table 5.** DENV and CHIKV co-positivity.

| S.No | Study ID | TaqMan™ CHIKV Ct value | CDC Trioplex CHIKV Ct value | TaqMan™ DENV Ct value | CDC Trioplex DENV Ct value | DENV Serology | CDC DENV 1-4 PCR |
| --- | --- | --- | --- | --- | --- | --- | --- |
| 1 | ZVT-35<sup>#</sup> | 26.22 | 31.50 | 36.22 | 32.09 | Not done | Negative |
| 2 | ZVT-09 | 12.38 | 16.39 | 35.98 | 0 | Negative | Negative |
| 3 | ZVT-25 | 17.82 | 22.33 | 36.14 | 0 | Negative | Negative |
| 4 | ZVT-26 | 19.43 | 23.71 | 36.89 | 0 | Negative | Negative |
| 5 | ZVT-52 | 23.09 | 26.98 | 35.93 | 0 | Negative | Negative |
| 6 | ZVT-53 | 22.29 | 26.08 | 36.97 | 0 | Negative | Negative |
| 7 | ZVT-54 | 21.27 | 25.48 | 35.92 | 0 | Negative | Negative |
| 8 | ZVT-28 | 21.29 | 24.83 | 35.17 | 0 | Negative | Negative |

<sup>#</sup>Concordant DENV and CHIKV results on TaqMan™ Arbovirus kit and CDC Trioplex.

**Table 6.** Performance of TaqMan™ Arbovirus kit for DENV.

| TaqMan™ Arbovirus Triplex | CDC Trioplex |  |  |  |
| --- | --- | --- | --- | --- |
|  |  | + | - | Total |
|  | + | 114 | 8 | 122 |
|  | - | 0 | 158 | 158 |
|  | Total | 114 | 166 | 280 |
| Sensitivity (CI 95%) | 100.00 (96.82 – 100.00) |  |  |  |
| Specificity (CI 95%) | 95.18 (90.73- 97.90) |  |  |  |
| Accuracy (%) | 97.14 |  |  |  |
| PPV (%) | 93.44 |  |  |  |
| NPV (%) | 100.00 |  |  |  |
| Kappa Coefficient (CI 95%) | 0.94 (0.90 - 0.98) |  |  |  |

Note. PPV, positive predictive value; NPV, negative predictive value.

When Ct values were compared, both assays showed a strong correlation for CHIKV regardless of Ct value (R^2^ = 0.9349) (Fig 3). However, the correlation for DENV was lower (R^2^ = 0.5927), with a stronger correlation observed between assays at lower Ct values compared to higher Ct values. Among the samples with concordant results on both assays, all except two DENV samples had Ct values below 35 on the TaqMan™ Arbovirus kit, with values of 36.22 and 35.51.

**Fig 3.**
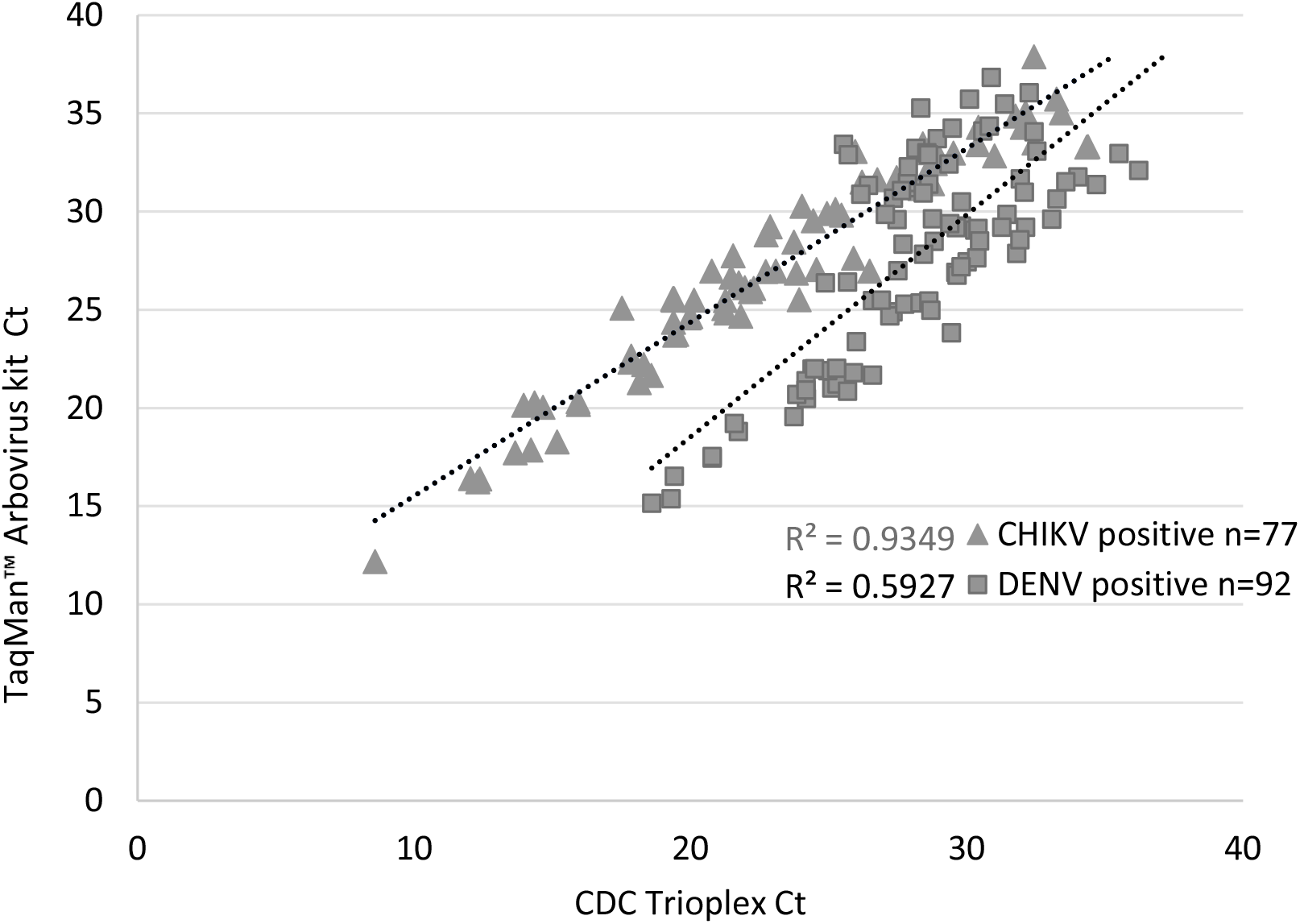
Ct values correlation between TaqMan™ Arbovirus kit and CDC Trioplex for CHIKV and DENV

### Costing

We undertook a costing exercise for the PCR kits with the Cost Accounting department of our institution. The CDC Trioplex PCR kit was provided free of cost for this study but required additional reagents such as SuperScript III Platinum One-step qRT-PCR kit and consumables (non-binding tubes, sterile filter tips, PCR plates, optical adhesive sealer, PCR plate applicator), personnel costs (cost to the institution), institutional overhead charges, and electricity. These additional charges were included for costing the PCR assays. The TaqMan™ Arbovirus kit on the other hand did not require any additional reagents and consumables as it was provided in the kit except for the institutional charges.

Based on this costing, the TaqMan™ Arbovirus kit assay costs approximately INR 1,340 per sample while the CDC Trioplex assay costs approximately INR 2,200 per sample.

## Discussion

Integrating multiplex PCR into routine testing for DENV, CHIKV, and ZIKV is crucial for enhancing pathogen detection while minimizing costs and constraints on human resources within existing health system capacities. This comparative analysis showed good sensitivity and specificity of the TaqMan™ Arbovirus kit with lower cost, lyophilized and ready-to-use reagents compared to the CDC Trioplex assay. The TaqMan™ Arbovirus kit demonstrated high sensitivity and specificity of 93.98% and 100% for CHIKV detection and 100% and 95.18% for DENV detection, respectively.

Both multiplex assays successfully detected circulating DENV serotypes. Among 122 DENV-positive samples subjected to CDC DENV-1-4 rRT-PCR, DENV2 and 3 were the most prevalent (n=40 each) serotypes positive by TaqMan™ Arbovirus kit followed by DENV1 (n=6). In comparison, out of 81 DENV-positive samples by CDC Trioplex, DENV3 was the most prevalent (n=39) followed by DENV2 (n=37) (**Supplement Table 4)**. The 5 samples that were serotype-negative were part of the 30 discordant samples listed in **Table 4**. All 86 samples positives by TaqMan™ Arbovirus kit and serotyped had Ct values below 35 Ct and the majority of the 81 positives by CDC Trioplex also had Ct values of below 35. The 5 samples missed by CDC Trioplex had Ct values ranging from 29-33 in TaqMan™ Arbovirus kit. The TaqMan™ Arbovirus kit showed more positives, likely due to the utilization of larger RNA input of 25 µl compared to the lower RNA input of 10 µl each used in the CDC Trioplex and CDC DENV 1-4 RT-PCR assays. The findings support an advantage for TaqMan™ Arbovirus kit, as it accurately identified the 5 DENV discordant samples further confirmed by positive DENV serology results (NS1/IgM/IgG).

Among 8 samples positive for both DENV and CHIKV using the TaqMan™ Arbovirus kit and negative for DENV on CDC Trioplex, only one was confirmed as a true co-infection **(Table 5)**. The remaining seven samples exhibited Ct values below 24 for CHIKV, indicative of high viral load and Ct values above 35 for DENV, implying weak positivity. The variation in RNA input volumes between the assays may have influenced the observed results. It is possible that while a DENV template of 25 µl may be beneficial for TaqMan™ Arbovirus kit, the opposite may be true for strong CHIKV virus infections, potentially resulting in non-specific/false positive DENV results.

A similar comparative study examining the performance of molecular tests for Zika virus detection highlighted the correlation between the volume of plasma used for extraction and the analytical sensitivity of the assays. The study determined the limit of detection (LOD) for the TaqMan™ Arbovirus kit as LOD_95_ 46.4 IU/mL, 95% CI: 23.5, 75.9, in contrast to the CDC Trioplex with LOD_95_ 457 IU/mL, 95% CI: 231, 749 and concluded that increased sensitivity was achieved by utilizing a larger plasma volume for the initial RNA extraction step. Specifically, the study employed a plasma volume of 200 µl for the CDC Trioplex and 300 µl for the TaqMan™ Arbovirus kit (24).

Our study findings also indicate the need to reevaluate the Ct value cutoff of 38 for the TaqMan™ Arbovirus kit. We observed only two DENV-positive samples with Ct values above 35 and no CHIKV sample. Similarly, all seven samples with co-infection, erroneously identified as DENV positive by the TaqMan™ Arbovirus kit, had Ct values above 35 for DENV. Lowering the Ct cutoff could potentially improve the assay performance for DENV detection without compromising the performance for CHIKV detection.

In addition to achieving high sensitivity and specificity, molecular tests should also demonstrate features such as affordability, short turnaround times, and robustness of reagents and consumables in terms of storage and usage (25). The TaqMan™ Arbovirus kit is conveniently provided in a user-friendly lyophilized format that eliminates the need for reagent preparation. By simply adding the sample template, this streamlined approach minimizes handling thereby reduces the risk of pre-analytical errors, with reagent contamination being the most common concern. Furthermore, this format speeds up the process from sample to result while costing approximately 40% less per sample than the comparator multiplex assay.

### Limitations of our study

The evaluation of the assay has certain limitations including the reliance on a single comparator and the lack of ZIKV-positive samples. Therefore, to strengthen this study, all DENV-positive samples were tested with the CDC DENV 1-4 rRT-PCR and the findings used to resolve the discordant results. Additionally, the absence of a specific limit of detection (LOD) assessment before testing presents another limitation. Addressing these challenges in future evaluations may contribute to a more comprehensive understanding of the assay’s performance and its potential utility in diagnostic settings.

## Conclusions

In conclusion, the TaqMan™ Arbovirus Triplex Kit (ZIKV/DENV/CHIKV) demonstrated high sensitivity and specificity (>93%) for the detection of circulating strains of CHIKV and DENV. Early detection may encourage prompt access to appropriate medical care that may potentially lower mortality and morbidity rates associated with severe infection. Given the simultaneous circulation of arboviruses viruses with similar clinical presentations, implementation of multiplex PCR testing for acute case detection and surveillance is essential. Accurate and timely detection enabled by multiplex PCR assays decreases unnecessary laboratory testing, optimizes use of laboratory resources and facilitates effective allocation of public health resources to control outbreaks.

## Supporting information

https://drive.google.com/drive/folders/1z_foz0kOy7tMUu3ixzv75NyqzPakAU2_?usp=sharing

## Data Availability

All data produced in the present study are available upon reasonable request to the authors. All data produced in the present work are contained in the manuscript.

## Data availability

The raw data supporting the conclusions of this article will be made available by the authors, without undue reservation.

## Ethics Statement

The Institutional Review Board of the Christian Medical College Vellore granted ethical approval.

## Supplementary material

Supplement Table 1a: CHIKV positive concordant samples

Supplement Table 1b: CHIKV negative concordant samples

Supplement Table 2: CHIKV discordant samples

Supplement Table 3a: DENV positive concordant samples

Supplement Table 3b: DENV negative concordant samples

Supplement Table 4: CDC DEN 1-4 rRT PCR positive results

## Authors contributions

SAH and OO: resources. AMA, SAH, SM and KP: investigation. KP: data curation. AMA, SM, and OO: conceptualization. AMA and SM: supervision. OO, KP and AMA: writing – original draft. AMA, SAH, SM, OO and KP: writing – review and editing. All authors contributed to the article and approved the submitted version.

## Funding

This work was supported by Thermo Fisher Scientific, who provided TaqMan™ Arbovirus Triplex Kit (ZIKV/DENV/CHIKV) intended for research purposes only.

## Declaration of Interests

OO is an employee of Thermo Fisher Scientific. All other authors declare no competing interest.

## Acknowledgements

The authors would like to thank the Centre of Disease Control and Prevention (CDC) for generously supplying the multiplex Trioplex and DENV 1-4 serotype rtRT-PCR. We thank all patients who provided samples for this evaluation. We also thank the staff of Thermo Fisher Scientific including Jelena Feenstra for contributions to the manuscript, Chakrapani Chatla who provided valuable insights during the study, and KC Ponnappa, for essential logistics support with the Thermo Fisher reagents used in this study.

## Notes

### Competing Interest Statement

The authors have declared no competing interest.

### Funding Statement

This study was funded by Invitrogen Bioservices India Pvt Ltd for consumables, reagents, PCR kit and article processing charge.

### Author Declarations

Institutional Review Board of Christian Medical College, Vellore gave ethical approval for this work. IRB Min. No. 14750 [DIAGNO] dated 27.07.2022

