## Supplementary material for "Performance evaluation of TaqMan^™^ Arbovirus Triplex Kit (ZIKV/DENV/CHIKV) for detection and differentiation of Dengue and Chikungunya viral RNA in serum samples of symptomatic patients": https://drive.google.com/drive/folders/1z_foz0kOy7tMUu3ixzv75NyqzPakAU2_?usp=sharing

**Supplement Tables:**

**Supplement Table 1a: CHIKV positive concordant samples**

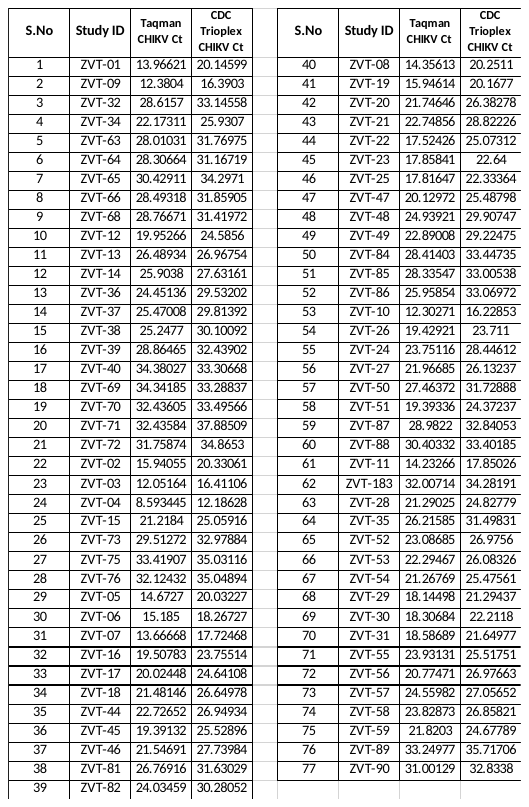

**Supplement Table 1b: CHIKV negative concordant samples**

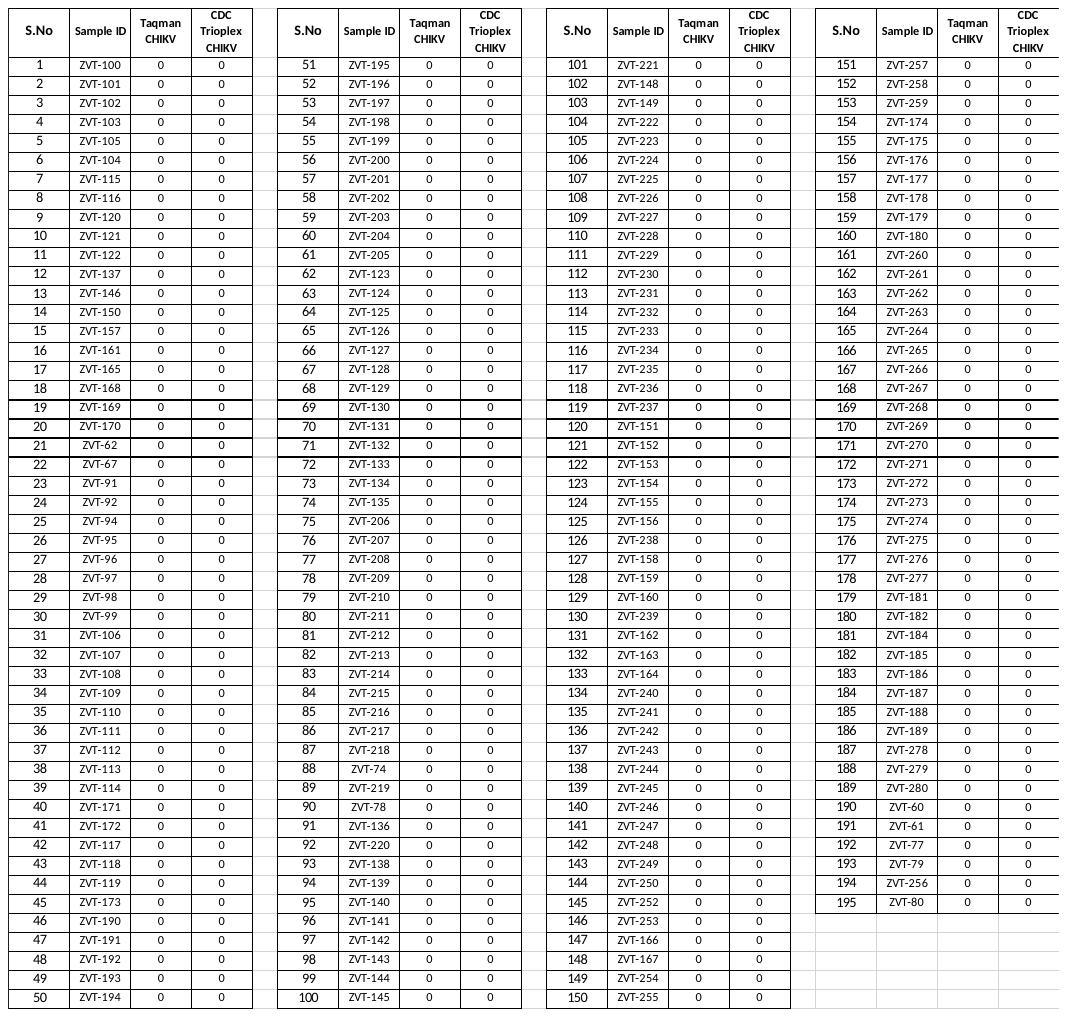

**Supplement Table 2: CHIKV discordant samples**

| **S.No** | **Study ID** | **TaqMan™ (CHIKV Ct value)** | **CDC Trioplex (CHIKV Ct value)** | **CHIKV IgM Antibodies** | **CHIKV Screening PCR** | **Consensus** | **Resolved by** |
| --- | --- | --- | --- | --- | --- | --- | --- |
| 1 | ZVT-33 | Negative | Positive (31.50) | Positive | 27.15 | Positive | CAT-2 & 3 |
| 2 | ZVT-41 | Negative | Positive (37.09) | Positive | 29.65 | Positive | CAT-2 & 3 |
| 3 | ZVT-42 | Negative | Positive (30.91) | Positive | 29.66 | Positive | CAT-2 & 3 |
| 4 | ZVT-43 | Negative | Positive (28.19) | Positive | 28.61 | Positive | CAT-2 & 3 |
| 5 | ZVT-147 | Negative | Positive (37.25) | Negative |  | Negative | CAT-2 & 3 |
| 6 | ZVT-83 | Negative | Positive (34.29) | Positive | 32.35 | Positive | CAT-2 & 3 |
| 7 | ZVT-251 | Negative | Positive (36.42) | Negative |  | Negative | CAT-2 & 3 |
| 8 | ZVT-93 | Positive (33.21) | Negative | Positive |  | Positive | CAT-2 |

**Supplement Table 3a: DENV positive concordant samples**

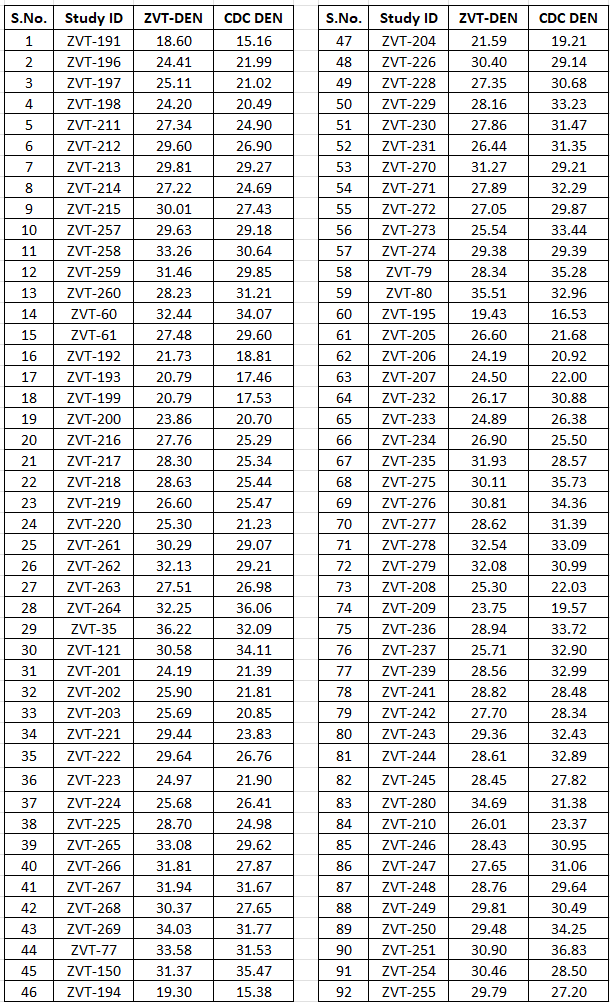

**Note.** ZVT, TaqMan™ Arbovirus kit PCR kit; CDC, CDC Trioplex assay

**Supplement Table 3b: DENV negative concordant samples**

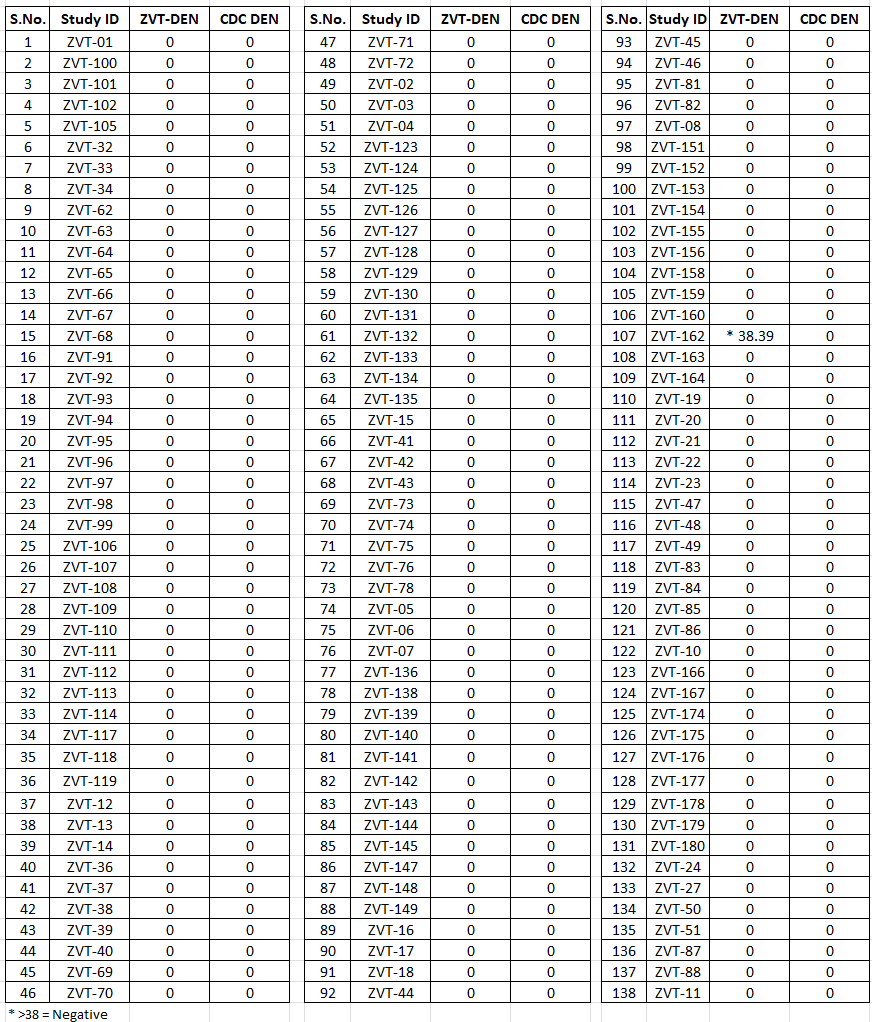

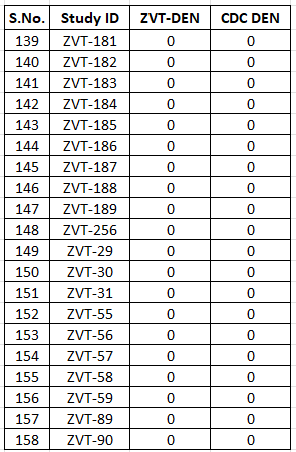

Note. ZVT, TaqMan™ Arbovirus kit PCR kit; CDC, CDC Trioplex assay

**Supplement Table 4: CDC DEN 1-4 rRT PCR positive results**

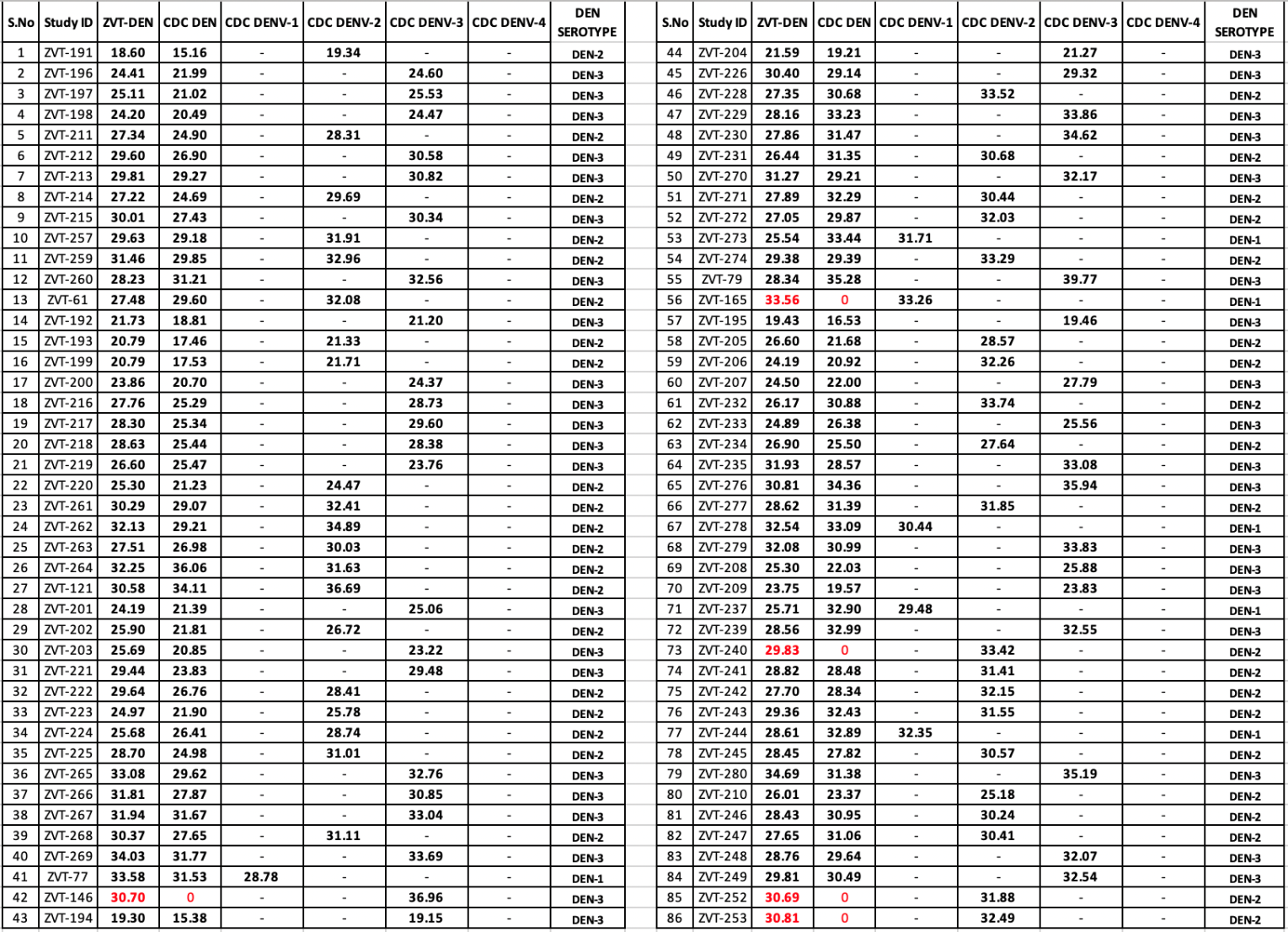

Note. ZVT, TaqMan™ Arbovirus kit PCR kit; CDC, CDC Trioplex assay; Highlighted in red are the TaqMan™ Arbovirus triplex (ZVT) DENV and CDC DEN 1-4 rRT PCR positives but negative by CDC Trioplex
